# DWI ASPECTS Can Detect Patients Who Require Puncture to Recanalization within 30 minutes for Large Vessel Occlusion

**DOI:** 10.1101/2023.03.23.23287670

**Authors:** Tomohide Yoshie, Toshihiro Ueda, Yasuhiro Hasegawa, Masataka Takeuchi, Masafumi Morimoto, Yoshifumi Tsuboi, Ryoo Yamamoto, Shogo Kaku, Junichi Ayabe, Takekazu Akiyama, Daisuke Yamamoto, Kentaro Mori, Hiroshi Kagami, Hidemichi Ito, Hidetaka Onodera, Yasuyuki Kaga, Haruki Ohtsubo, Kentaro Tatsuno, Noriko Usuki, Satoshi Takaishi, Yoshihisa Yamano, the K-NET Registry Investigators

**Affiliations:** St. Marianna University Toyoko Hospital; National Cerebral and Cardiovascular Center; St. Marianna University School of Medicine; Seisho Hospital; Yokohama Shintoshi Neurosurgical Hospital; Kawasakisaiwai Hospital; Yokohama Brain and Spine Center; Neurosurgical East Yokohama Hospital; Yokosuka Kyosai Hospital; Akiyama Neurosurgical Hospital; Kitasato University Hospital; Yokohama Sakae Kyosai Hospital; Saiseikai Yokohamashi Tobu Hospital; St.Marianna University Yokohama Seibu Hospital

**Keywords:** DWI ASPECTS, Puncture-to-Recanalization time, Mechanical thrombectomy, Acute ischemic stroke, Large vessel occlusion

## Abstract

**BACKGROUND:** Faster recanalization for acute large vessel occlusion is associated with better clinical outcome. The aim of this study was to identify patient characteristics that require puncture-to-recanalization (P-R) time within 30 min for good clinical outcome.

**METHODS:** We evaluated the patients from K-NET Registry, which is a prospective, multicenter, observational registry of acute ischemic stroke patients. The present analysis included patients who met the following criteria: (1) treated with endovascular therapy for ICA or MCA M1 occlusion; and (2) achieved successful recanalization (mTICI 2b or more). Patients were divided into subgroups according to pre-treatment patients’ characteristics; DWI ASPECTS, age, NIHSS, occlusion site and symptom recognition to puncture time. In each subgroup, the frequency of good clinical outcomes was compared between P-R time <30min and >30min. We entered interaction terms into the models to evaluate the correlation between each patient characteristic and P-R time.

**RESULTS:** 1053 patients were included in this study. Overall, P-R <30min was significantly associated with good outcome. Univariate analysis in each subgroup revealed a significant association between P-R <30min and good outcomes in the patients with DWI ASPECTS ≤6, age >85 and NIHSS ≥16. In multivariable analysis, NIHSS, age, symptom recognition to puncture time, DWI ASPECTS and P-R <30 minutes were significant independent predictors of good outcomes; however, only DWI ASPECTS had interaction terms with P-R <30 minutes. Multivariable analysis showed P-R <30min was an independent predictor for good outcome in patients with DWI ASPECTS ≤6, whereas P-R<30min was not in patients with DWI ≥7.

**CONCLUSIONS:** P-R<30 min is a predictor of good clinical outcome, however, the effect depends on DWI ASPECTS. A target P-R time of less than 30 minutes is appropriate for patients with DWI ASPECTS scores of 0-6, whereas 30 minutes or longer is reasonable for patients with DWI ASPECTS 7 or higher.

## Introduction

Time to reperfusion is a critical factor in achieving good clinical outcomes for endovascular thrombectomy in acute ischemic stroke due to large vessel occlusion. Studies have shown that shorter time to reperfusion is associated with better clinical.(1, 2, 3) Stroke systems of care should be developed to ensure that mechanical thrombectomy-eligible patients receive treatment as quickly as possible. However, it is not clear whether all thrombectomy-eligible patients require fast recanalization. Moreover, if time to reperfusion is only associated with good outcomes in certain patients, it is important to identify the background of those who require fast recanalization.

The ideal time for each step of the process, including puncture-recanalization (P-R) time, is also unclear. The recommended target P-R time is 60 minutes.(4) Studies have shown that the rate of favorable outcomes is significantly higher when procedural time is less than 60 minutes.(5, 6). However, the P-R time in previous randomized controlled trials was shorter than 60 minutes, with a median time of 24 minutes in the SWIFT PRIME trial(7) and 44 minutes in the HERMES collaboration (1). Another suggested target P-R time is 30 minutes. The Society of Neuro-Interventional Surgery has proposed stroke process time metrics that recommend door-to-puncture time within 60 minutes and door-to-recanalization time within 90 minutes, requiring an ideal P-R time of within 30 minutes.(8) Although P-R time within 30 minutes may be an appropriate target time, it can be difficult to achieve for some cases. It would be useful for the endovascular treatment team to know which patients require P-R time within 30 minutes.

In this study, we investigated the association between clinical outcomes and P-R time in a large-scale multicenter registry. The aim of this study was to identify the background of patients who require P-R time within 30 minutes for good clinical outcomes.

## Methods

### Study Population

The data that support the findings of this study are available from the corresponding author upon reasonable request. The Kanagawa Intravenous and Endovascular Treatment of Acute Ischemic Stroke registry (K-NET Registry) is a prospective, multicenter, observational study of patients with acute ischemic stroke who received intravenous tPA therapy and/or endovascular therapy for acute ischemic stroke in 40 stroke centers in Kanagawa Prefecture, Japan. The protocol and primary results of the K-NET registry have been previously published.(9) The inclusion criteria for the K-NET registry were patients who received intravenous tPA therapy for acute ischemic stroke and/or had an intention to perform endovascular therapy for large vessel occlusion. There were no exclusion criteria.

The present study is based on the data from patients included in the K-NET registry between January 2018 and June 2020. The inclusion criteria for the present analysis were as follows: (1) ICA or MCA M1 occlusion; (2) treated with endovascular therapy; and (3) achieved successful recanalization, defined as an mTICI score of 2b or more.

In the K-NET Registry, basic information such as patients’ background, imaging findings, functional outcomes, procedure details, and time of treatment, including P-R time, were collected. All data were entered into a web-based electronic case report system by the treating physicians. The data underwent standardized quality checks to control for consistency, plausibility, and completeness. Data that had unresolved queries were excluded.

### Ethics Approval

The study protocol of the K-NET registry was approved by the Ethics Committee of St. Marianna University School of Medicine (approval no. 3757). According to local regulations, further approval was obtained from local institutional ethics committees or institutional review boards. Written informed consent was obtained from the patient or the surrogate. The information required for registration was obtained from routine clinical practice.

### Imaging and Endovascular Treatment

CT or MRI were used for diagnostic imaging at admission. The imaging methods used to evaluate thrombectomy-capable patients (MRI, CTA, or CTP) were determined by each institution. The evaluation of ischemic lesions in MRI was made with a Diffusion-weighted Imaging–Alberta Stroke Program Early Computed Tomography Scores (DWI-ASPECTS) of 0-11. The EVT method was determined by neuro-interventionalists at each institution based on mechanical thrombectomy, thrombus aspiration, and intra-arterial thrombolysis. There were no restrictions on the selection of the EVT devices. The recanalization grade was measured by the modified Thrombolysis in Cerebral Infarction (mTICI) scale. Well-experienced neurologists, neurosurgeons, or neuroradiologists in each institution evaluated DWI ASPECTS and mTICI scale.

### Outcome

The outcome was measured by the mRS score at 90 days after the stroke onset. The primary outcome was a good outcome, defined as mRS 0-2. As this study also included patients with pre-stroke mRS 3-5, no decrease in mRS score was also defined as a good outcome, but only in patients with pre-stroke mRS 3-5, in concordance with the definitions used in previous reports.(10)

### Statistical analysis

Baseline characteristics were analyzed using standard statistics. Mean and standard deviation (SD) were used for continuous variables, and median with interquartile range (IQR) were used for ordinal variables. Proportions were used for categorical variables. To evaluate the effect of P-R time on outcome, patients were grouped into subgroups according to their background characteristics: age (<70, 70-85, >85), NIHSS (≤5, 6-10, 11-15, 16-20, ≥21), occlusion site (ICA, MCA), DWI ASPECTS (≤6, 7-8, 9-10, no MRI), and symptom recognition to puncture time (≤1.5h, 1.5-3h, 3-4.5h, 4.5-6h, ≥6). In each subgroup, the frequency of good clinical outcomes was compared between P-R time <30min and >30min using χ2 tests. Multivariable stepwise logistic regression models were then used to determine predictors of good outcome, with patients’ background including P-R time as variables. As DWI ASPECTS was not determined in the no MRI group, these patients were excluded from the multivariable analysis. To evaluate the correlation between each background characteristic and P-R time, interaction terms for all variables were also entered into the models. Odds ratios, confidence intervals, and corresponding p-values were derived, with a p-value < 0.05 considered significant. Statistical analyses were conducted using SPSS (Version 9.4, IBM, USA) and Stata 17 (Stata Corporation, College Station, TX).

## Results

Between January 2018 and June 2020, 2316 patients were included in the K-NET registry, and 1165 patients underwent endovascular therapy for ICA or MCA M1 occlusion. Of these, 112 patients with incomplete recanalization (mTICI 0, n=37; mTICI 1, n=20; mTICI 2a, n=55) were excluded. Finally, 1053 patients were included in this analysis (Supplemental Figure 1).

Patients’ backgrounds are presented in Table 1. The mean age was 75.0±12.1 years. The number of patients aged <70, 70-85, and >85 years was 272 (25.8%), 583 (55.3%), and 198 (18.8%), respectively. The median NIHSS was 19 (IQR 14-24), and the number of patients with NIHSS scores of ≤5, 6-10, 11-15, 16-20, and ≥21 was 62 (5.8%), 109 (10.3%), 170 (16.1%), 269 (25.5%), and 443 (42.0%), respectively. ICA occlusion was present in 441 (41.8%) patients, and MCA M1 occlusion was present in 612 (58.1%) patients. The median DWI ASPECTS was 7 (IQR 6-9), and DWI ASPECTS scores of ≤6, 7-8, and 9-10 were observed in 304 (28.8%), 232 (22.0%), and 233 (22.1%) patients, respectively. Two hundred eighty-four patients (26.9%) did not undergo MRI before endovascular therapy. The median time from symptom recognition to puncture was 139 min (IQR 104-196 min), and the number of patients with symptom recognition to puncture times of ≤1.5h, 1.5-3h, 3-4.5h, 4.5-6h, and ≥6h was 153 (14.5%), 583 (55.3%), 185 (17.5%), 48 (4.5%), and 84 (4.5%), respectively.

**Table 1.**
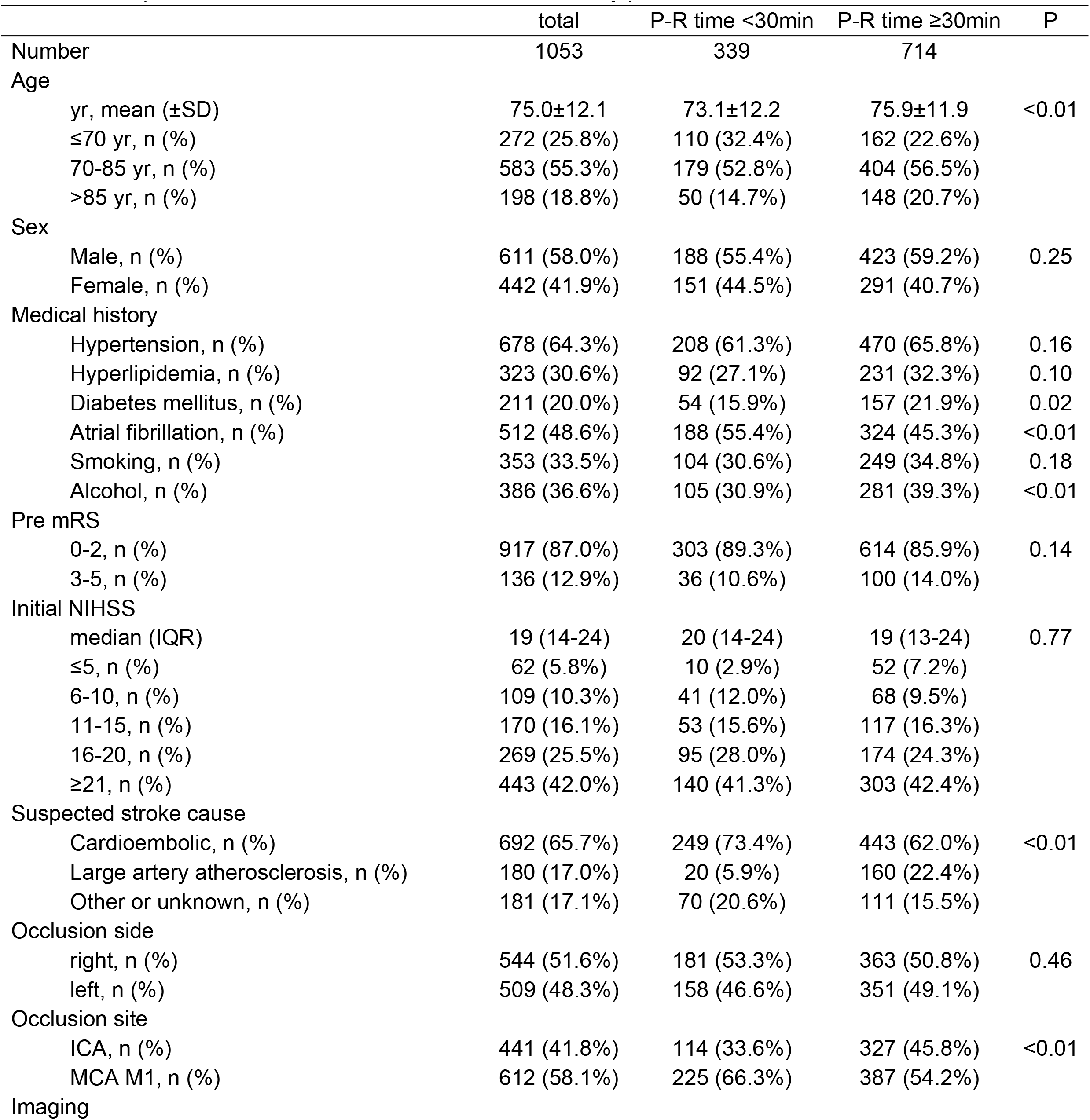
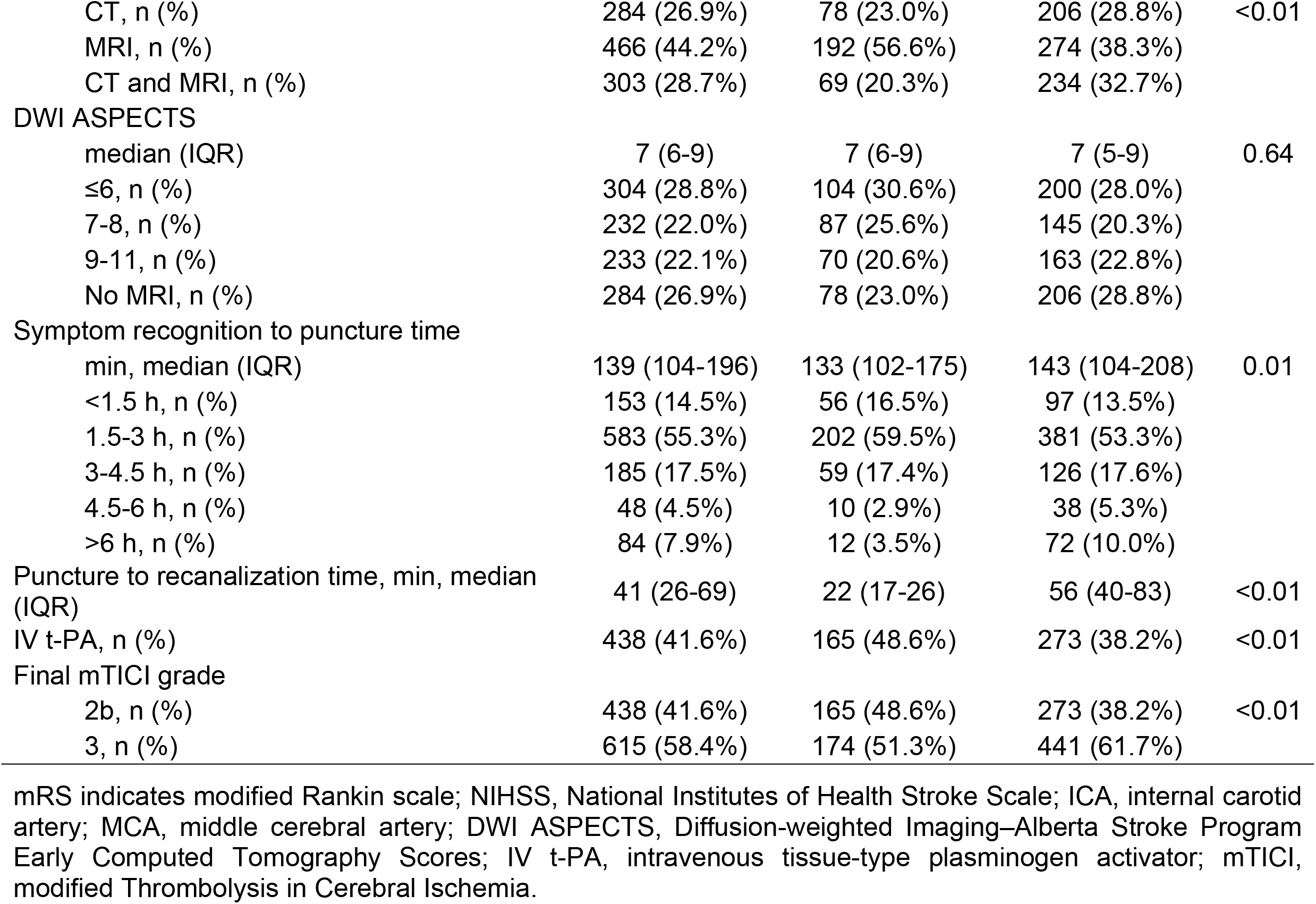
Comparison of Baseline Characteristics of Patients by puncture-recanalization time

|  | total | P-R time <30min | P-R time ≥30min | P |
| --- | --- | --- | --- | --- |
| Number | 1053 | 339 | 714 |  |
| Age |  |  |  |  |
| yr, mean (±SD) | 75.0±12.1 | 73.1±12.2 | 75.9±11.9 | <0.01 |
| ≤70 yr, n (%) | 272 (25.8%) | 110 (32.4%) | 162 (22.6%) |  |
| 70-85 yr, n (%) | 583 (55.3%) | 179 (52.8%) | 404 (56.5%) |  |
| >85 yr, n (%) | 198 (18.8%) | 50 (14.7%) | 148 (20.7%) |  |
| Sex |  |  |  |  |
| Male, n (%) | 611 (58.0%) | 188 (55.4%) | 423 (59.2%) | 0.25 |
| Female, n (%) | 442 (41.9%) | 151 (44.5%) | 291 (40.7%) |  |
| Medical history |  |  |  |  |
| Hypertension, n (%) | 678 (64.3%) | 208 (61.3%) | 470 (65.8%) | 0.16 |
| Hyperlipidemia, n (%) | 323 (30.6%) | 92 (27.1%) | 231 (32.3%) | 0.10 |
| Diabetes mellitus, n (%) | 211 (20.0%) | 54 (15.9%) | 157 (21.9%) | 0.02 |
| Atrial fibrillation, n (%) | 512 (48.6%) | 188 (55.4%) | 324 (45.3%) | <0.01 |
| Smoking, n (%) | 353 (33.5%) | 104 (30.6%) | 249 (34.8%) | 0.18 |
| Alcohol, n (%) | 386 (36.6%) | 105 (30.9%) | 281 (39.3%) | <0.01 |
| Pre mRS |  |  |  |  |
| 0-2, n (%) | 917 (87.0%) | 303 (89.3%) | 614 (85.9%) | 0.14 |
| 3-5, n (%) | 136 (12.9%) | 36 (10.6%) | 100 (14.0%) |  |
| Initial NIHSS |  |  |  |  |
| median (IQR) | 19 (14-24) | 20 (14-24) | 19 (13-24) | 0.77 |
| ≤5, n (%) | 62 (5.8%) | 10 (2.9%) | 52 (7.2%) |  |
| 6-10, n (%) | 109 (10.3%) | 41 (12.0%) | 68 (9.5%) |  |
| 11-15, n (%) | 170 (16.1%) | 53 (15.6%) | 117 (16.3%) |  |
| 16-20, n (%) | 269 (25.5%) | 95 (28.0%) | 174 (24.3%) |  |
| ≥21, n (%) | 443 (42.0%) | 140 (41.3%) | 303 (42.4%) |  |
| Suspected stroke cause |  |  |  |  |
| Cardioembolic, n (%) | 692 (65.7%) | 249 (73.4%) | 443 (62.0%) | <0.01 |
| Large artery atherosclerosis, n (%) | 180 (17.0%) | 20 (5.9%) | 160 (22.4%) |  |
| Other or unknown, n (%) | 181 (17.1%) | 70 (20.6%) | 111 (15.5%) |  |
| Occlusion side |  |  |  |  |
| right, n (%) | 544 (51.6%) | 181 (53.3%) | 363 (50.8%) | 0.46 |
| left, n (%) | 509 (48.3%) | 158 (46.6%) | 351 (49.1%) |  |
| Occlusion site |  |  |  |  |
| ICA, n (%) | 441 (41.8%) | 114 (33.6%) | 327 (45.8%) | <0.01 |
| MCA M1, n (%) | 612 (58.1%) | 225 (66.3%) | 387 (54.2%) |  |
| Imaging |  |  |  |  |
| CT, n (%) | 284 (26.9%) | 78 (23.0%) | 206 (28.8%) | <0.01 |
| MRI, n (%) | 466 (44.2%) | 192 (56.6%) | 274 (38.3%) |  |
| CT and MRI, n (%) | 303 (28.7%) | 69 (20.3%) | 234 (32.7%) |  |
| DWI ASPECTS |  |  |  |  |
| median (IQR) | 7 (6-9) | 7 (6-9) | 7 (5-9) | 0.64 |
| ≤6, n (%) | 304 (28.8%) | 104 (30.6%) | 200 (28.0%) |  |
| 7-8, n (%) | 232 (22.0%) | 87 (25.6%) | 145 (20.3%) |  |
| 9-11, n (%) | 233 (22.1%) | 70 (20.6%) | 163 (22.8%) |  |
| No MRI, n (%) | 284 (26.9%) | 78 (23.0%) | 206 (28.8%) |  |
| Symptom recognition to puncture time |  |  |  |  |
| min, median (IQR) | 139 (104-196) | 133 (102-175) | 143 (104-208) | 0.01 |
| <1.5 h, n (%) | 153 (14.5%) | 56 (16.5%) | 97 (13.5%) |  |
| 1.5-3 h, n (%) | 583 (55.3%) | 202 (59.5%) | 381 (53.3%) |  |
| 3-4.5 h, n (%) | 185 (17.5%) | 59 (17.4%) | 126 (17.6%) |  |
| 4.5-6 h, n (%) | 48 (4.5%) | 10 (2.9%) | 38 (5.3%) |  |
| >6 h, n (%) | 84 (7.9%) | 12 (3.5%) | 72 (10.0%) |  |
| Puncture to recanalization time, min, median (IQR) | 41 (26-69) | 22 (17-26) | 56 (40-83) | <0.01 |
| IV t-PA, n (%) | 438 (41.6%) | 165 (48.6%) | 273 (38.2%) | <0.01 |
| Final mTICI grade |  |  |  |  |
| 2b, n (%) | 438 (41.6%) | 165 (48.6%) | 273 (38.2%) | <0.01 |
| 3, n (%) | 615 (58.4%) | 174 (51.3%) | 441 (61.7%) |  |

mRS indicates modified Rankin scale; NIHSS, National Institutes of Health Stroke Scale; ICA, internal carotid artery; MCA, middle cerebral artery; DWI ASPECTS, Diffusion-weighted Imaging–Alberta Stroke Program Early Computed Tomography Scores; IV t-PA, intravenous tissue-type plasminogen activator; mTICI, modified Thrombolysis in Cerebral Ischemia.

The median P-R time was 44 min (IQR 26-69 min). P-R times of <30 min and ≥30 min were observed in 339 (32.1%) and 714 (67.8%) patients, respectively. In the group with P-R times of <30 min, the patients were significantly younger, had a lower incidence of diabetes mellitus, higher incidence of atrial fibrillation and M1 occlusion, and shorter symptom recognition to puncture times. This group had a significantly higher rate of using IV t-PA and a lower rate of TICI 3 recanalization.

### Outcome and Puncture to Recanalization Time in Subgroups

Figure 1 and Supplemental Table 1 show the comparison of good outcomes between the P-R time <30 minutes and ≥30 minutes in subgroups based on patients’ characteristics. Overall, at 90 days after thrombectomy, 196 (57.8%) patients in the P-R time <30 minutes group had a good outcome compared to 331 (46.3%) in the P-R time ≥30 minutes group (P<0.01). In the subgroup analysis, P-R time <30 minutes was associated with good outcomes in subgroups consisting of patients with DWI ASPECTS ≤6 (good clinical outcome, n (%), 51 (49.0%) in P-R time <30min vs 52 (26.0%) in P-R time ≥30min, p<0.01), age >85 years old (26 (52.0%) vs 41 (27.7%), p<0.01), ICA occlusion (63 (55.2%) vs 127 (38.8%), p<0.01), NIHSS 16-20 (55 (57.8%) vs 78 (44.8%), p=0.04), NIHSS ≥21 (65 (46.4%) vs 97 (46.1%), p<0.01), symptom-recognition-puncture-time <1.5 hours (41 (73.2%) vs 43 (44.3%), p<0.01) and symptom-recognition-puncture-time 1.5-3 hours (114 (56.4%) vs 43 (45.6%), p=0.02). In contrast, there was no significant differences in rates of good clinical outcome between P-R time <30 minutes and ≥30 minutes in subgroups with DWI ASPECTS ≥7, age ≤85 years old, MCA M1 occlusion, NIHSS ≤15, or symptom-recognition-puncture-time ≥3 hours.

**Figure 1.**
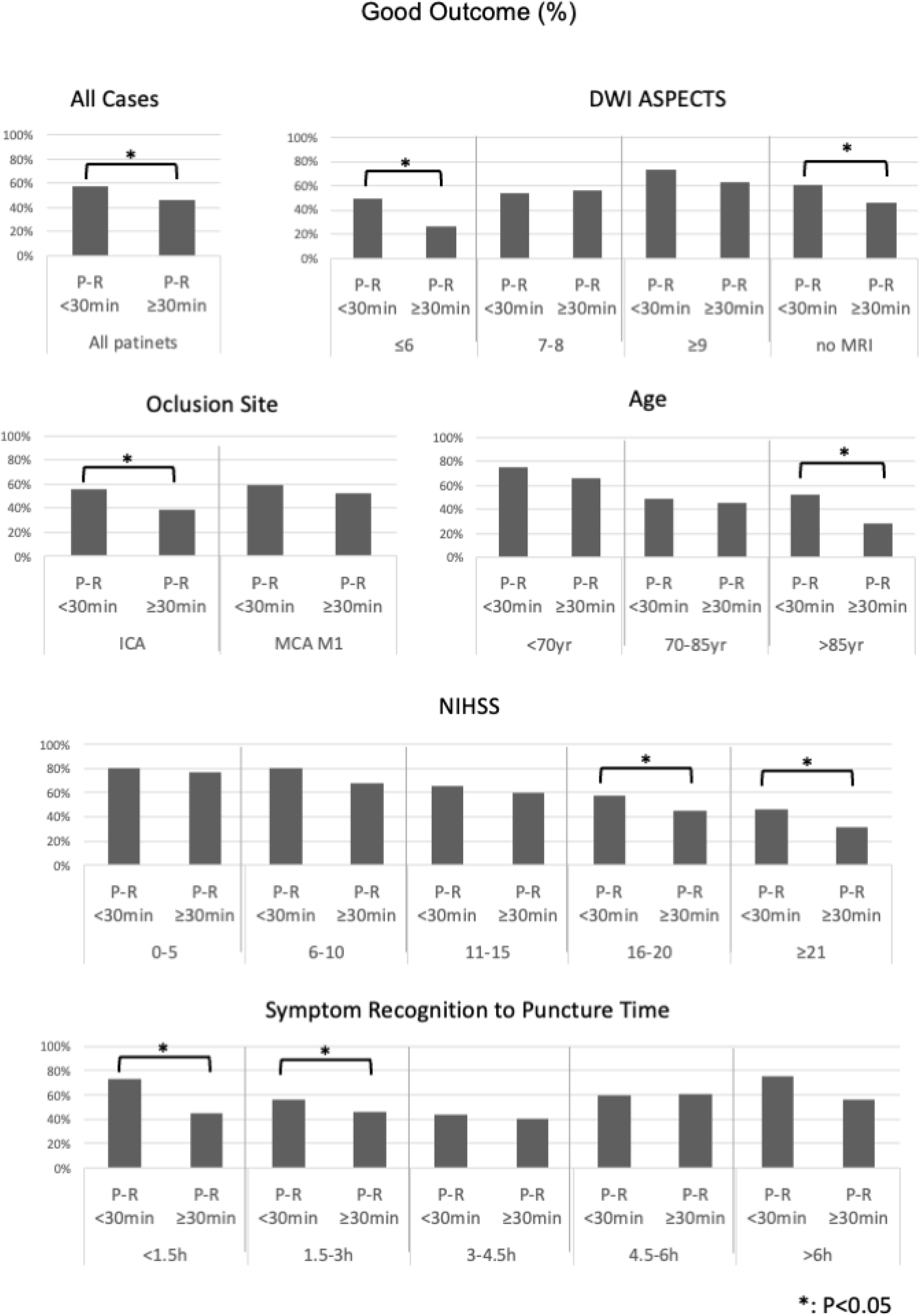
Comparing probability of good clinical outcome between puncture-to-recanalization time less than 30 min and more than 30min in each subgroup according to baseline characteristics of patients. Asterisk (*) indicates P<0.05. P-R time indicates puncture-to-recanalization time; DWI ASPECTS, Diffusion-weighted Imaging–Alberta Stroke Program Early Computed Tomography Scores; ICA, internal carotid artery; MCA, middle cerebral artery; NIHSS, National Institutes of Health Stroke Scale

### Multivariable Regression Analysis with Interaction Terms

Table 2 shows the results of a multivariable logistic regression analysis with interaction terms for predicting good outcome at 90 days. NIHSS (P<0.01), age (P<0.01), and symptom recognition to puncture time (P<0.01) were significant independent predictors of good outcome. DWI ASPECTS (P<0.01) and P-R <30min (P=0.02) were also significant independent predictors of good outcome. However, only DWI ASPECTS had an interaction term with P-R <30min for predicting good clinical outcome (P<0.01). The patients’ and treatment characteristics, other than DWI ASPECTS, did not have statistically significant interaction terms with P-R <30min.

**Table 2.**
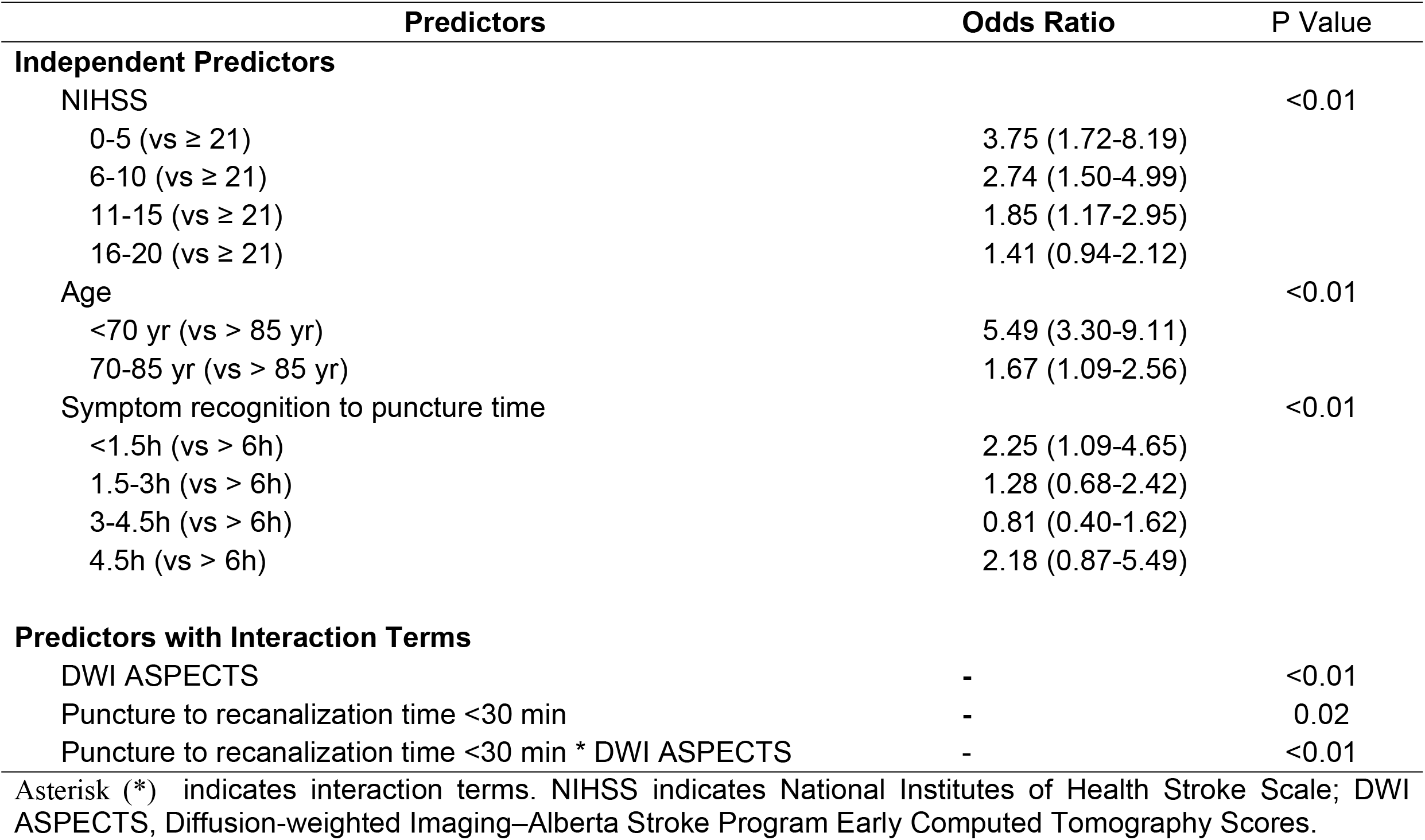
Multivariable Logistic Regression Analysis with Interaction Terms for Predictors of Good Outcome at 90 days

| Predictors | Odds Ratio | P Value |
| --- | --- | --- |
| **Independent Predictors** |  |  |
| NIHSS |  | <0.01 |
| 0-5 (vs ≥ 21) | 3.75 (1.72-8.19) |  |
| 6-10 (vs ≥ 21) | 2.74 (1.50-4.99) |  |
| 11-15 (vs ≥ 21) | 1.85 (1.17-2.95) |  |
| 16-20 (vs ≥ 21) | 1.41 (0.94-2.12) |  |
| Age |  | <0.01 |
| <70 yr (vs > 85 yr) | 5.49 (3.30-9.11) |  |
| 70-85 yr (vs > 85 yr) | 1.67 (1.09-2.56) |  |
| Symptom recognition to puncture time |  | <0.01 |
| <1.5h (vs > 6h) | 2.25 (1.09-4.65) |  |
| 1.5-3h (vs > 6h) | 1.28 (0.68-2.42) |  |
| 3-4.5h (vs > 6h) | 0.81 (0.40-1.62) |  |
| 4.5h (vs > 6h) | 2.18 (0.87-5.49) |  |
| **Predictors with Interaction Terms** |  |  |
| DWI ASPECTS | - | <0.01 |
| Puncture to recanalization time <30 min | - | 0.02 |
| Puncture to recanalization time <30 min * DWI ASPECTS | - | <0.01 |

Asterisk (\*) indicates interaction terms. NIHSS indicates National Institutes of Health Stroke Scale; DWI ASPECTS, Diffusion-weighted Imaging–Alberta Stroke Program Early Computed Tomography Scores.

Since only DWI ASPECTS was a significant independent predictor for good outcome with an interaction with P-R<30min, we conducted a multivariable logistic regression analysis in subgroups based on DWI ASPECTS. We found that P-R time <30min was a significant independent predictor of good clinical outcome in patients with DWI ASPECTS ≤6 (Odds ratio 2.58 (95% CI 1.54-4.34), P<0.01). Age and NIHSS were significant independent predictors of good outcome in the DWI ASPECTS 7-8 group, and age was a significant independent predictor in the DWI ASPECTS ≥ 9 group. However, P-R<30min was not a significant independent predictor of good outcome in these groups (Table 3).

**Table 3.** Multivariable Logistic Regression Analysis for Good Outcome in subgroups by DWI ASPECTS

| Predictors for good outcome |  | Odds Ratio (95% CI) | P Value |
| --- | --- | --- | --- |
| **DWI ASPECTS≤6** |  |  |  |
| Puncture to recanalization time <30min | <30min (vs ≥ 30min) | 2.58 (1.54-4.34) | <0.01 |
| Age | <70yr (vs >85yr) | 4.15 (1.90-9.06) | <0.01 |
|  | 70-85yr (vs >85yr) | 1.39 (0.66-2.90) |  |
| **DWI ASPECTS 7-8** |  |  |  |
| Age | <70yr (vs >85yr) | 8.91 (3.45-3.02) | <0.01 |
|  | 70-85yr (vs >85yr) | 1.30 (0.63-2.68) |  |
| NIHSS | ≤5 (vs ≥ 21) | 15.54 (1.85-30.3) | <0.01 |
|  | 6-10 (vs ≥ 21) | 4.46 (1.28-5.51) |  |
|  | 11-15 (vs ≥ 21) | 1.70 (0.76-3.80) |  |
|  | 16-20 (vs ≥ 21) | 1.05 (0.52-2.13) |  |
| **DWI ASPECTS ≥ 9** |  |  |  |
| Age | <70yr (vs >85yr) | 4.80 (1.94-1.86) | <0.01 |
|  | 70-85yr (vs >85yr) | 2.31 (1.16-4.58) |  |

DWI ASPECTS indicates Diffusion-weighted Imaging–Alberta Stroke Program Early Computed Tomography Scores; NIHSS, National Institutes of Health Stroke Scale

## Discussion

In the present study, we found that a P-R time of less than 30 minutes is a predictor of good clinical outcome. However, the impact of P-R time on outcome depends on DWI ASPECTS. P-R time less than 30 minutes is an independent predictor of good clinical outcome only in patients with DWI ASPECTS ≤6. No association was found between outcome and P-R time of less than 30 minutes in patients with DWI ASPECTS ≥7. Our study suggests that the target P-R time for a good outcome is within 30 minutes in patients with DWI ASPECTS ≤6. In contrast, a target time of 30-60 minutes or more is optimal for good clinical outcome in patients with DWI ASPECTS ≥7.

In this study, a duration of 30 minutes was chosen as the target time from puncture to recanalization. However, achieving recanalization within this timeframe is often difficult in clinical practice. In our study, the median puncture to recanalization time was 42 minutes, and 67.7% of cases took over 30 minutes. In a recent registry, the median puncture to recanalization time was reported to be 37-70 minutes.(11, 12, 13) Puncture to recanalization time tends to be longer in patients with certain conditions, such as old age (14), tandem occlusion and tortuous arteries. Additionally, less experienced endovascular operators may find it challenging to achieve recanalization within 30 minutes, as procedural duration is associated with the operator’s experience. (15) Our findings suggest that patients with low DWI ASPECTS should be treated by experienced endovascular operators, as they may have better success rates in achieving timely recanalization.

A previous study showed that patients with a procedure time within 30 minutes had lower rates of unfavorable outcomes at discharge compared with patients with procedure times over 30 minutes.(16) Another study using ADAPT thrombectomy technique showed that the likelihood of achieving a good outcome was higher in patients who achieved puncture to recanalization within 35 minutes than over 35 minutes.(14) Several studies showed that a procedure time within 30 minutes is a predictor of good clinical outcome, however, these studies did not assess subgroups according to patients’ backgrounds. There are a few studies that showed the interaction between ASPECTS and onset-reperfusion time. Todo et al. assessed the interaction between CT-ASPECTS and onset-reperfusion time.(17) They suggested that patients with lower ASPECTS should be targeted for a shorter onset-reperfusion time than patients with higher ASPECTS. In the current study, we suggest that the ideal procedure time depends on DWI ASPECTS.

Our study showed that the ideal puncture to recanalization time is 30 minutes for endovascular treatment for patients with ASPECTS 0-6. A recent randomized controlled trial showed that patients with large ischemic core had better functional outcomes with endovascular therapy than with medical care alone. (18, 19, 20) However, the rate of good clinical outcomes (mRS 0-2) was low, ranging from 14.0% to 30.0%. A previous meta-analysis that assessed the impact of endovascular therapy in patients with pretreatment ASPECTS scores of 0–6 showed that only the onset-to-recanalization time was significantly associated with functional independence.(21) To improve the outcome of endovascular therapy for patients with large ischemic cores, it is necessary to achieve a puncture-to-recanalization time of less than 30 minutes, as suggested by our results.

There are several reasons why the association between puncture-to-recanalization time and outcome depends on DWI ASPECTS scores. First, patients with low DWI ASPECTS scores have severe tissue damage, and delayed reperfusion of such lesions can result in hemorrhagic transformation and reperfusion injury. Another reason is that patients with low DWI ASPECTS scores have poor collaterals and low cerebral blood flow, which are the main determinants of ASPECTS decay.(22) A previous study showed that optimal cerebral blood flow thresholds associated with follow-up infarction depend on time to reperfusion.(23) Indeed, ASPECTS at presentation could predict a fast infarct growth rate in patients receiving endovascular therapy. (24)

Strengths of our study include the large number of patients who underwent MRI before thrombectomy. There are several limitations to our study. First, it is a retrospective analysis of a single-arm prospective registry and does not include a control arm. Potential sources of bias were identified in our retrospective analysis of the registry study, including selection bias and measurement bias. To address these potential sources of bias, a standardized protocol for data collection was developed to minimize measurement bias. In K-NET registry, all data were entered into a web-based electronic case report system by the treating physicians. The data underwent standardized quality checks to control for consistency, plausibility, and completeness. Also, we adjusted for potential confounding variables, such as age, NIHSS and time from symptom-recognition to arriving hospital, in our analysis to control for confounding bias. We used multivariable regression analysis to adjust for these potential confounding variables and to identify independent predictors of outcome. Second, MRI was not performed in all patients, 26.9% patients underwent CTA or CTP before endovascular therapy instead of MRI. Third, there was no central imaging core laboratory for diagnostic imaging in our study. DWI ASPECTS and mTICI scores were evaluated in each institution. However, these imaging data were evaluated by well experienced neurologists, neurosurgeons, or neuroradiologists in each institution, and all data underwent a standardized central data quality check. Additionally, DWI has high inter-rater agreement compared to CT.(25) Finally, since our study included patients with pre-stroke mRS scores of 2 or more, we defined a good clinical outcome as an mRS score of 0-2 or no decrease in the mRS score at 90 days after stroke. The common definition of a good clinical outcome is an mRS score of 0-2, as many previous studies excluded patients with pre-stroke mRS scores of 2 or more.

In conclusion, DWI ASPECTS scores can identify patients who require a puncture-to-recanalization time of less than 30 minutes to achieve a good outcome. Our study suggests that a target time from puncture to recanalization of less than 30 minutes is appropriate for patients with DWI ASPECTS scores of 0-6, whereas a time of 30-60 minutes or longer is reasonable for patients with DWI ASPECTS scores of 7 or higher.

## Data Availability

We stated as follows in the methods section: The data that support the findings of this study are available from the corresponding author upon reasonable request.

## Acknowledgments

We would like to thank the K-NET Registry investigators. We are grateful to Ms. Tomomi Shibuya and Mr. Satoshi Muta (Department of Practical Management of Medical Information, St. Marianna University School of Medicine) for collecting and checking data.

## Supplemental Material

Table S1

Figure S1

